# Perceptions and willingness of patients and caregivers on the utilization of patient-generated health data: a cross-sectional survey

**DOI:** 10.1101/2024.10.31.24316537

**Authors:** Ye-Eun Park, Sang Sook Beck, Yura Lee

## Abstract

**Background:** Patient-generated health data (PGHD) are increasingly recognized for their potential to complement traditional healthcare data by offering continuous monitoring and enhanced patient management. Despite the growing interest in PGHD however, the perceptions and willingness of patients and caregivers to utilize this information in a clinical setting remain underexplored.

**Objective:** This study aimed to assess the perceptions, expectations, and concerns of patients and caregivers regarding the clinical utilization of PGHD. The influence of demographic factors such as age and health status on these perceptions was also explored.

**Methods:** A cross-sectional survey was conducted of 400 participants, including both patients and caregivers. The survey collected data on attitudes towards PGHD, experiences with health information management, and the willingness to share PGHD for clinical and secondary purposes. Statistical analysis was used to identify significant differences in perceptions based on demographics and health-related roles.

**Results:** The analysis revealed significant variations in attitudes towards PGHD that were based on the participants’ health-related roles, age, and gender. Older patients and male caregivers exhibited higher concerns about data privacy and security, while younger participants showed greater enthusiasm for using PGHD in managing their health. These findings highlighted diverse needs and expectations across different demographic groups.

**Conclusion:** Consideration of demographics and role-based differences is very important when designing and implementing PGHD systems. Tailored approaches that address specific concerns and expectations can enhance the acceptance and effectiveness of PGHD in clinical practice, ultimately fostering more patient-centered care.

## Introduction

Patient-generated health data (PGHD) are created, recorded, or gathered by or from patients, family members or other caregivers to help address a health concern.(1) PGHD have been increasingly recognized in contemporary healthcare, primarily due to the unique benefits provided that extend beyond traditional healthcare data generated within medical institutions.(2) PGHD offer insights from a different perspective, aiding patients through continuous care, monitoring, and high-risk management. As the scope of PGHD includes everything from Patient-Reported Outcome Measures (PROMs) in classical clinical trials to various lifestyle choices related to health, this has emerged as a means to resolve the information blackout between medical visits.(3)

As a large axis of data for more precise medical services, PGHD are expected to contribute to a lowering the burden of medical costs and improvements in healthcare through a focus on prevention and high-risk management rather than on the services provided at hospitals and clinics.(4–6) In addition to continuity of care, PGHD are important in that they can better reflect real-world outcomes of clinical interventions by highlighting the patient’s environment and health-related choices based on personal tendencies.(3, 7) PGHD provide greater transparency on the impacts of the care from medical institutions and can thereby improve the benefits of these interventions, such as improving the health outcomes of patients who actively use PGHD or through the co-production of applications between healthcare providers, caregivers and patients to ensure continuity of care.(8–10)

The use of PGHD is not without challenges and concerns, however. The ambiguous nature of the boundaries of PGHD poses significant issues regarding data protection and privacy.(11–13) The inherent difficulty in safeguarding this information, coupled with challenges in its integration, also makes it vulnerable to breaches.(14) Furthermore, the diverse needs associated with PGHD, given the broad categories involved, complicates its effective utilization.(3) These concerns highlight the necessity to adequately address privacy risks and ensure robust protection mechanisms to foster trust and widespread acceptance of PGHD among patients and caregivers.

Given the aforementioned benefits and challenges, it will be imperative going forward to understand the perceptions of patients and caregivers regarding the use of PGHD. To achieve the meaningful and patient-centered utilization of these data, it is crucial to comprehend the views and attitudes of the primary stakeholders—the patients themselves. The sharing of clinically relevant information, including PGHD, is closely linked to patient engagement, which acts as a catalyst for improved treatment adherence, patient safety, and overall health outcomes. Thus, this study aimed to investigate the awareness and perceptions of PGHD among patients and caregivers, which are essential for driving patient participation in PGHD and achieving its full potential in healthcare.

## Methods

The primary purpose of this survey study was to investigate patients’ expectations and concerns about PGHD. We conducted a structured survey with the aim of examining the perceptions of patients and caregivers regarding the clinical utilization and secondary use of PGHD. Additionally, we sought to explore how PGHD is generated differently based on the characteristics of patients and their caregivers (such as age and disease classification) and to understand how they consumed the services associated with their PGHD.

### Questionnaire and Survey Design

The questionnaire used in the study survey was designed to obtain data on how patients and caregivers primarily manage their health information and in what format, how they engage with Personal Health Record (PHR) services affiliated with healthcare institutions, and their perceptions regarding the secondary utilization of PGHD. The development process for this questionnaire involved discussions among the authors of this study, which included a medical regulatory/legal expert (SSB), and medical informatics experts (YL, YEP). The survey underwent further modifications and enhancements through consultation with experts in healthcare institution-affiliated PGHD research (SY), a health IT industry professional (SYS), and a specialist in medical information-related laws and ethics (KY).(6, 7, 13, 15–18)

The categories used in the finalized questionnaire were broadly as follows:

(A) General information, including type of disease, health-related (care-related) role of respondent (patient/caregiver)
(B) Experience with health information management services with or without connection to institutions’ Electronic Health Record (EHR)
(C) Willingness to consent to the primary (medical) purposes of using the PGHD within the medical institution,
(D) Willingness to consent to secondary (research; commercial) uses of the PGHD,
(E) Participatory processes that the respondents consider most important or are most curious about.

The response formats for each survey item were measured according to the nature of the questionnaire. These formats included the 4-point Likert scale (ranging from 1=Strongly Disagree to 4=Strongly Agree), Yes/No responses, single-choice selections, multiple-choice selections, and open-ended responses.

### Respondent enrolment

A survey was conducted from 6 to 12 November 2023 targeting unspecified patients and caregivers who may utilize or have the potential to utilize PGHD. Participants were recruited through various patient communities including the Korean Leukemia Patient Association, Korean Type 1 Diabetes Patient Association, Korean Kidney Cancer Patient Association, Korean Neuroendocrine Tumor Patient Association, Korean GIST (Gastrointestinal Stromal Tumor) Patient Association, Korean Congenital Heart Disease Patient Association, Cancer Patient Solidarity, and Korean Psoriasis Association. We recruited these participants by posting an announcement on public online patient communities, which included a link and a QR code to access the survey. We ensured that participants who provided consent to utilize their survey responses for research could respond to the structured questionnaire. Our aim was to recruit over 100 respondents each for chronic diseases, malignant diseases, and other diseases, because we considered the disease type to be most crucial feature for distinguishing subgroups.

### Statistical Analysis

Following the completion of the survey, for a more detailed comparison of characteristics among survey respondents, we categorized these subjects into disease groups (congenital/genetic conditions, traumatic conditions, malignant conditions, chronic conditions, rare conditions, and others), health-related role of the respondents i.e. the patients themselves (Pt); caregivers of minor patients (CG-minor); caregivers of patients aged 70 or older or those unable to attend medical appointments alone (CG-elder); caregivers of adult patients not falling into the previous categories (CG-adult). We also stratified the participants by age group (20s, 30s, 40s, 50s and above).

We divided the survey participants into the following four groups to examine and compare their basic characteristics: Pt, CG-minor, CG-elder, and CG-adult, aiming to discern differences among responses through technical analysis of the survey data. We utilized the Kruskal-Wallis test in particular to verify the significance of response differences among three or more subgroups for categorical variables. Additionally, responses using a 4-point Likert scale, considered as continuous variables, were analyzed using one-way ANOVA. Prior to conducting the one-way ANOVA, we performed the Bartlett test to assess the homogeneity of variances across groups. In this analysis, response differences were considered statistically significant if the obtained p-value was less than 0.05. Data analysis was conducted using R (version 4.3.0).

## Results

The final number of participants amounted to 400 after verifying and removing 4 duplicate responses. In analyzing the distribution of gender across the health-related roles of the respondent groups, as shown in **Table 1**, it was found that the CG-elder group had the highest proportion of males (43.2%). With regard to the age distribution, the Pt group exhibited the highest percentages in both the 20s and over 50 categories which was statistically significant (*P* < 0.01). Regarding primary health issues, the Pt, CG-minor, and CG-elder groups predominantly reported chronic diseases (Pt: 40.3%, CG-minor: 60.9%, CG-elder: 44.6%), whereas the CG-adult group showed the highest prevalence of malignant diseases (56.6%). This discrepancy was statistically significant with a p-value below 0.01.

**Table 1.**
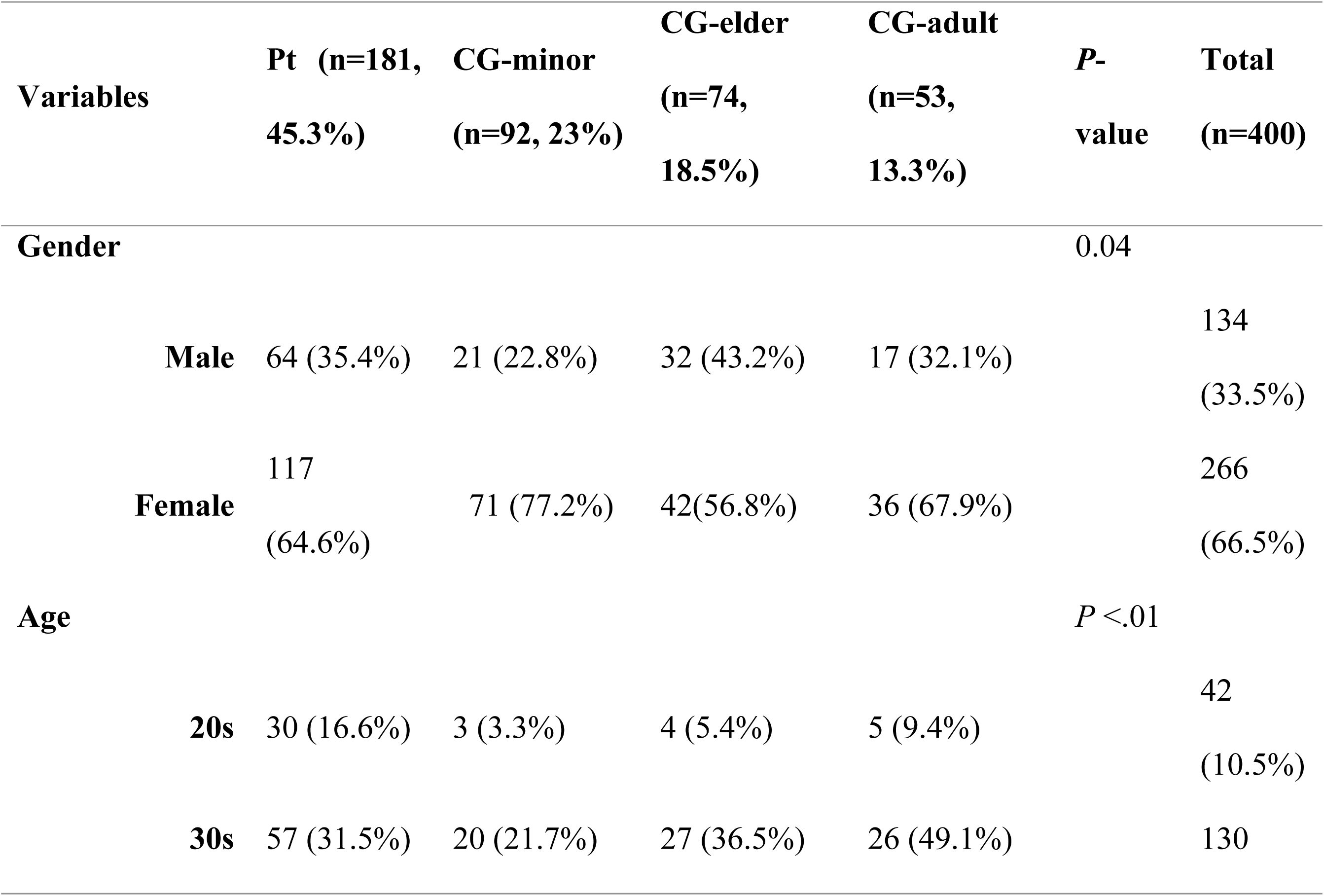
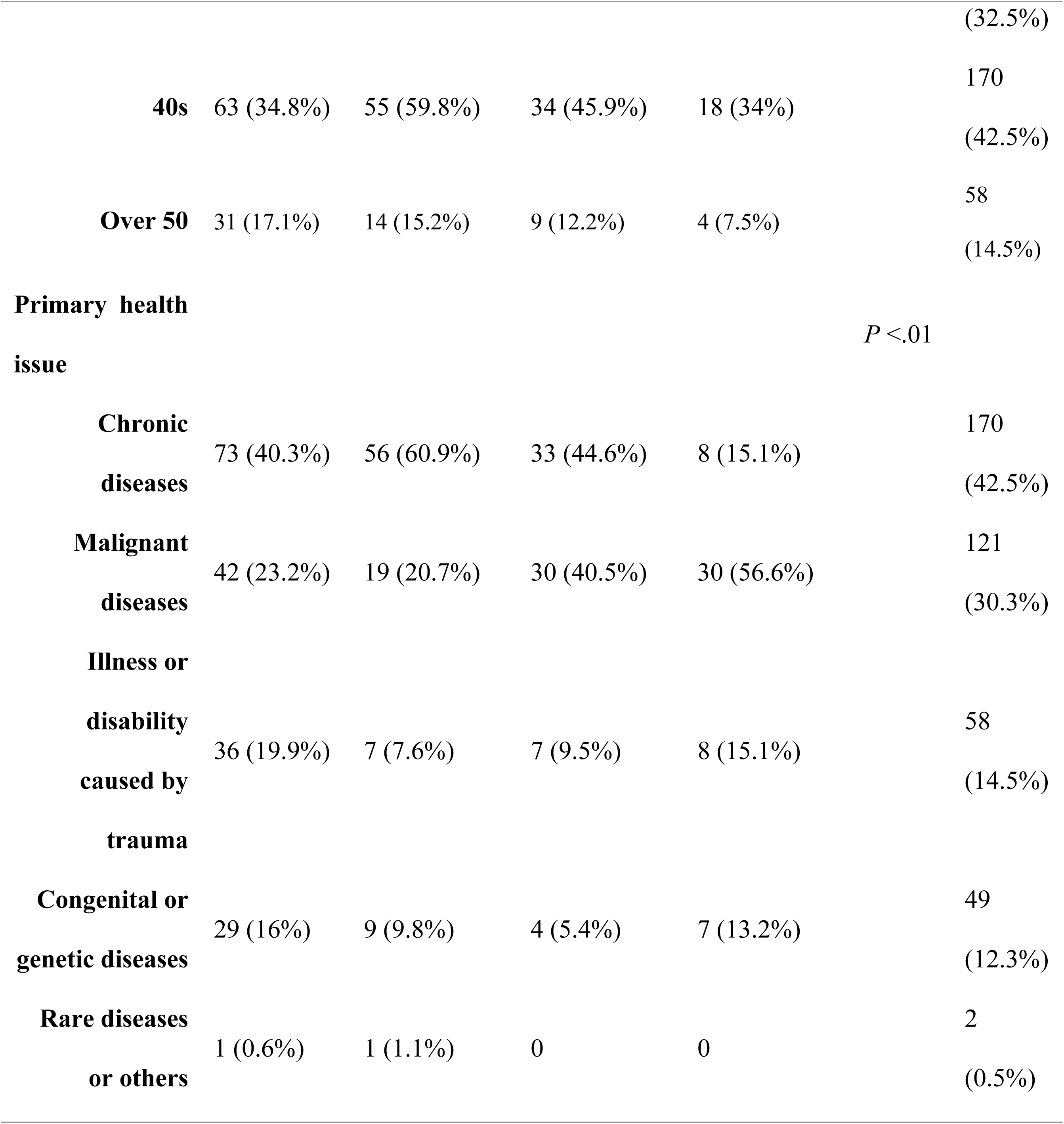
Demographic and Health Characteristic Comparisons Across the Health-related Roles of the Respondent Groups (Survey category A1-A4)

An analysis was conducted on all survey items for each group except for the open-ended responses, and significant differences were identified (**Tables 2 and 3**). Detailed analysis of the survey items for each group can be found in the **Supporting Information (S1-3 Table)**.

**Table 2.** Analysis of Survey Item Significance Across the Participant Category Groups: Experience with Health Information Management Services.

| No. | Questions | Disease<br>Group | Health-Related Role | Age<br>Group |
| --- | --- | --- | --- | --- |
| A5 | What methods do you usually use to record or manage your/your family's main health problems? | * | - | - |
| B1. | Have you ever used commercially available or downloadable apps from the app store (or a built-in app on your smartphone, such as Samsung Health or Apple Health) or a health information measuring device (such as a smartwatch) to measure or record your blood pressure, blood sugar, step counts, physical activity, and heart rate? | *** | *** | ** |
| B2. | Have you ever used a patient app provided by your healthcare organization? | * | * | - |
| B2a. | → (Y) Have you ever managed or searched for health information for yourself and your family using a patient app? | ** | - | - |
| B2b. | → (N) If you answered no, would you use a health management app service if your medical institution were to offer it? | *** | * | ** |

Significant relationships are denoted by \* ( $P < .05$ ), \*\* ( $P < .01$ ), and \*\*\* ( $P < .001$ )

**Table 3.**
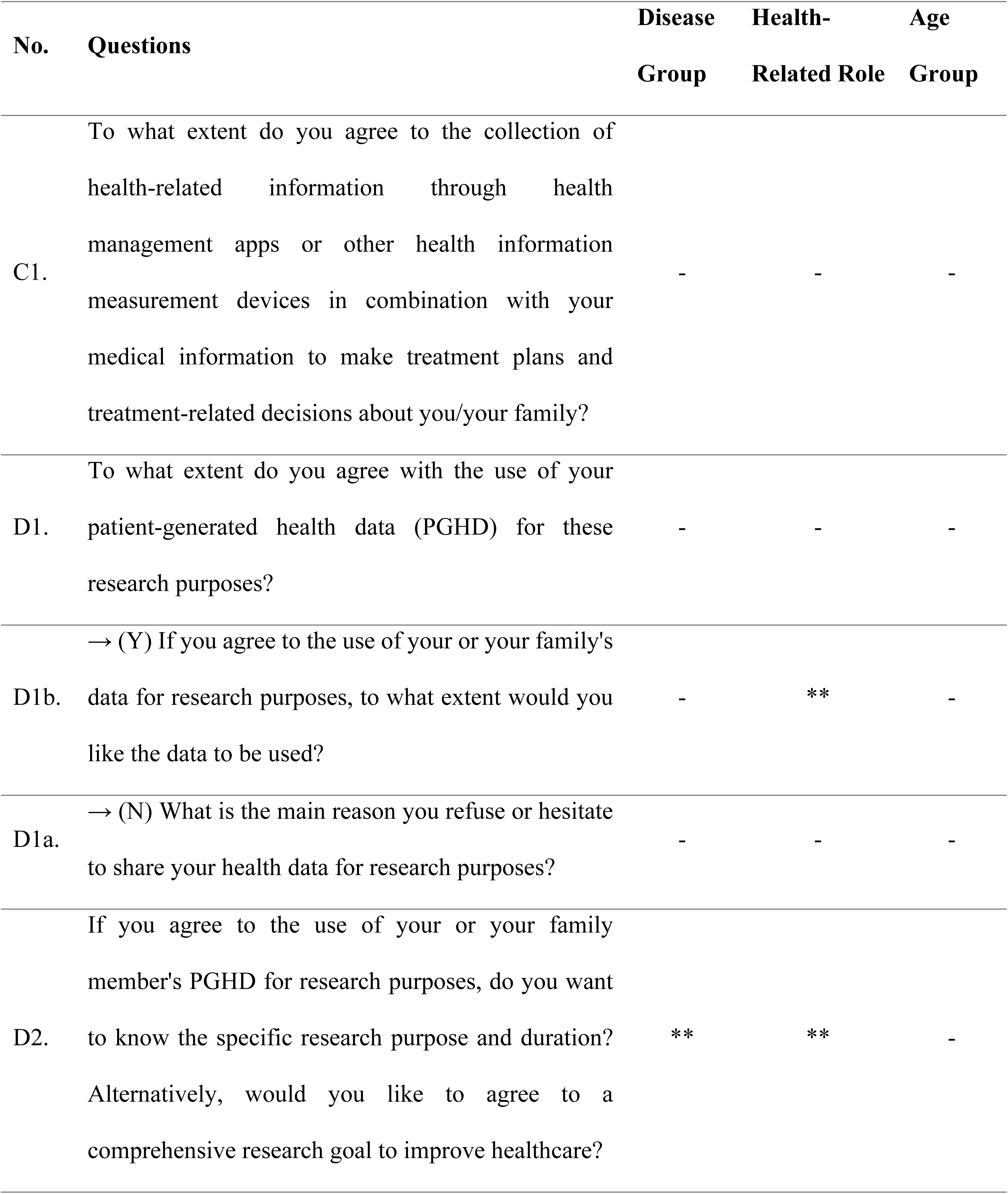
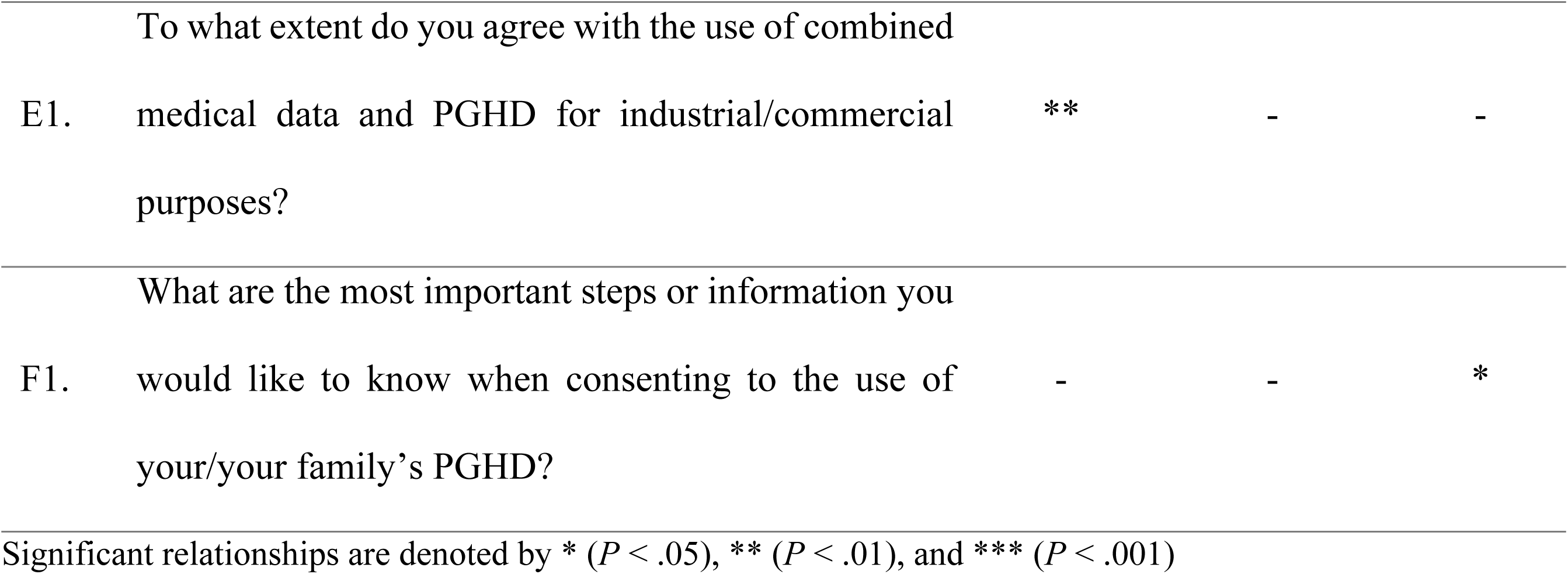
Analysis of Survey Item Significance Across Participant Category Groups: Attitude Toward the Use of PGHD.

### Experience with Health Information Management Services: Significant Differences in Survey Items Across Disease-, Health-related Role, and Age-groups

Regarding the methods for recording or managing health issues, analysis based on respondent diseases revealed that those with a primary traumatic condition showed the highest tendency to "not usually record or manage" at 24.1% (n=14/58), compared to other groups (Congenital/Genetic: 8.2%, n=4/49; Malignant: 13.2%, n=16/121; Chronic: 11.2%, n=19/170; Rare/Others: 0%, n=0/2). “Basic functions of a cell phone, smartphone, or a tablet such as photo album or note-taking apps” was the most frequent method to record or manage health information (151/400, 37.8%), except for respondents with chronic disease, 41.8% (n=71/170) of whom reported using “apps dedicated to disease/health management on a smartphone or tablet”.

The question asking whether respondents had experienced using commercial PHR apps (i.e., smartphone applications or devices) revealed distinct patterns based on disease type, health-related role, and age. In terms of disease type, respondents with chronic conditions reported the highest level of user experience with commercial PHR apps, with 78.8% (n=134/170) answering "Yes," compared to 37.9% (n=22/58) in the traumatic conditions group. Among all subjects who responded yes (n=247), the largest number of people indicated (multiple selection) that they measure physical activity (n=178), followed by blood sugar (n=114), heart rate (n=106), and blood pressure (n=85). Regarding health-related roles, the CG-minor group had the highest experience with PHR apps at 75% (n=69/92), followed by the Pt group at 66.9% (n=121/181). When considering age, the 20s age group had the lowest experience at 52.4% (n=22/42), whereas the 40s age group had the highest at 70.6% (n=120/170).

When asked about the duration of PHR app use among those who had experience, 63.6% of the traumatic condition group reported less than 6 months of use (n=14/22), the only disease group where a majority of the respondents reported this duration. In terms responses to this question stratified by age, respondents in their 30s were the only group to approach a majority reporting usage of less than 6 months (49.2%; n=34/69). With regard to frequency of updating categorized by health-related roles, both the Pt and CG-minor groups reported daily recording in a majority of cases (Pt: 52.1%, n=63/121; CG-minor: 63.8%, n=44/69). When the respondents indicated why they stopped using PGHD apps, the most common reasons given were ‘Because it was bothersome’ (43.1%, 69/160), ‘Because it was difficult to use’ (30.0%, 48/160), and ‘Because it was not helpful for my health management’ (22.5%, 36/160).

For the usage of patient apps provided by medical institutions (tethered PHR), respondents with malignant conditions reported the highest usage at 24% (n=29/121) compared to other disease groups (Congenital/Genetic: 16.3%, n=8/49; Traumatic: 27%, n=10/58; Chronic: 11.8%, n=20/170; Rare/Others: 0%, n=0/2). Regarding health-related roles, the Pt group responded ‘yes’ at the highest rate (20.4%, 37/181), followed by CG-minor (18.5%, 17/92). About two-thirds of the tethered PHR users responded that they have accessed health information through tethered PHR (65.1%, 41/63), and regardless of the respondent groups, the most commonly selected benefit of doing this was ‘It was helpful in the self-management of health and diseases’ (63.4%, 26/41). Regarding the willingness to use tethered PHR, analysis by health-related role showed that the CG-minor respondents had the highest intention to use at 86.8% (n=65/75). When classified by age group, the intention to use was lowest among those in their 20s at 60.5% (n=23/38). Conversely, respondents in their 40s and older expressed strong intentions to use, with rates exceeding 70% (40s: 81.9%, n=118/170; over 50: 73.9%, n=34/58).

### Attitude Toward the Use of PGHD: Significant Differences in Survey Items Across Disease-, Health-related Role, and Age-groups

Regarding the use of PGHD, respondents responded positively in the following order, regardless of group: clinical use (89.3%, 357/400), research (84.8%, 339/400), and commercial/industrial use (74.8%, 299/400) (**Figure 1**). In terms of the reasons for not agreeing to the use of this information for research purposes, the largest number of respondents (47.5%, 29/61) chose “Concerns regarding potential damage or disadvantage due to personal information leakage.” This was also the reason given as the issue of most concern with regard to commercial/industrial use (62.7%, 63/102) regardless of disease, health-related role, or age. Regarding the consent for data utilization, in terms of health-related roles, CG-minor respondents had the highest willingness for comprehensive data utilization including identifiable data at 58% (n=47/92). Conversely, the CG-adult group showed a tendency towards preferring anonymous data inclusion at 72.1% (n=31/53).

**Figure 1.**
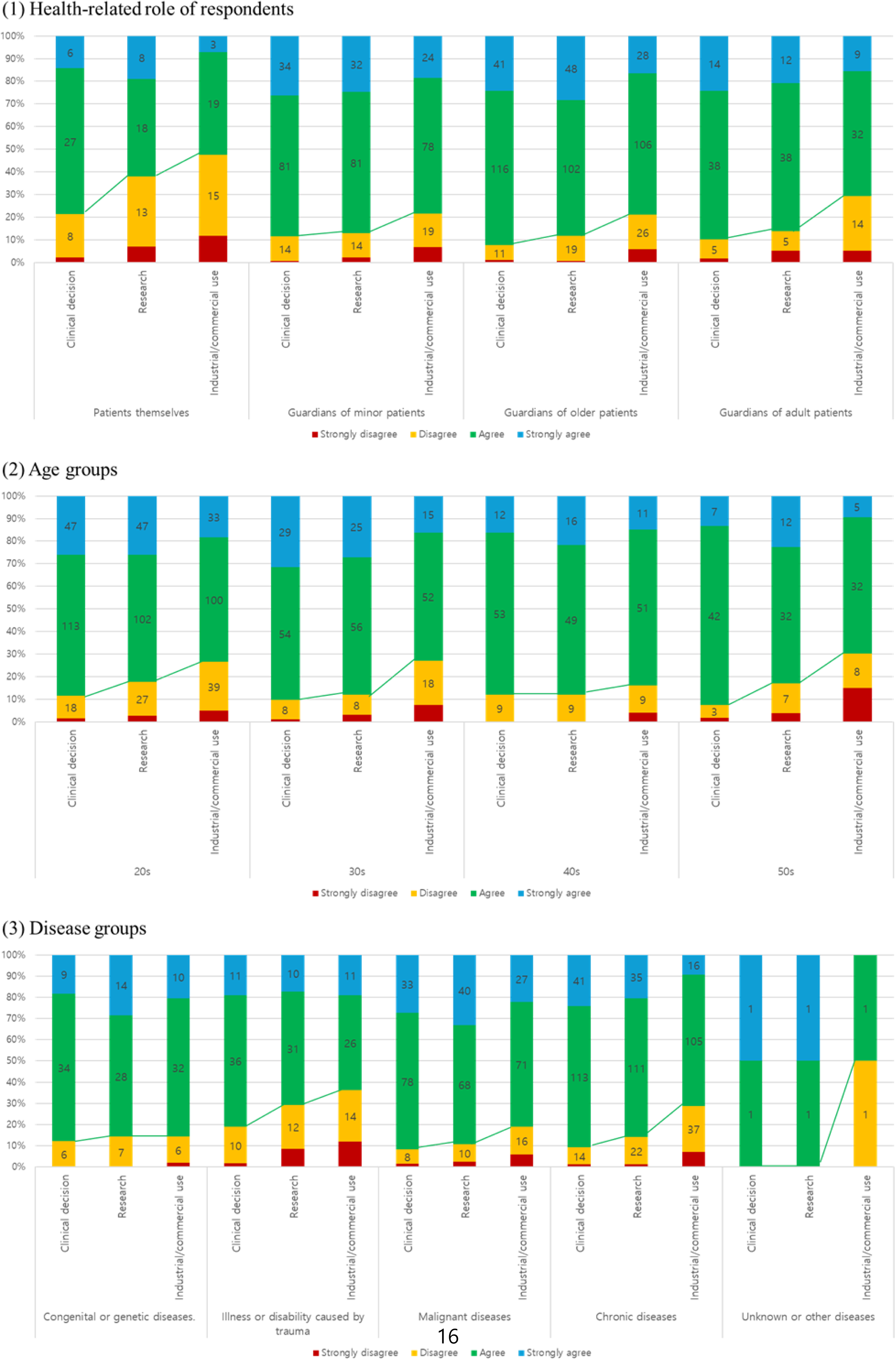
Attitude toward the use of PGHD in accordance with the utilization purpose across the Disease-, Health-related Role, and Age-groups

When assessing consent for the use of PGHD for research purposes, respondents were asked whether they would prefer to give agreement to certain studies on an individual basis (Consent by specific research purpose) or provide a blanket agreement that would require no further permission requests (Comprehensive consent). Among the disease classifications, respondents with malignant conditions were the only group where a majority (52.9%, n=64/121) opted for Comprehensive consent. Regarding health-related roles, the CG-elder participants showed the highest inclination towards Comprehensive consent at 55.4% (n=41/74).

### Responses on the Consent for Secondary Use

**Table 4** presents the top 5 most frequent responses regarding the reasons for choosing specific consent by specific research purpose or comprehensive consent in response to the question, “If you agree to the use of your or your family member’s PGHD for research purposes, do you want to know the specific research purpose and duration? Alternatively, would you like to agree to a comprehensive research goal to improve healthcare?” For respondents who chose specific consent by specific research purpose, the reasons for their choice were as follows, listed in the order of frequency: “Want to know specific research purposes, contents, and duration,” “Want to know how personal information is utilized in research,” and “Concerns about privacy breach, misuse, etc.” On the other hand, for respondents who selected comprehensive consent, the reasons for their choice were as follows, listed in order of frequency: “For convenience,” “Think it will be helpful for research,” and “For time-saving due to busyness.”

**Table 4.**
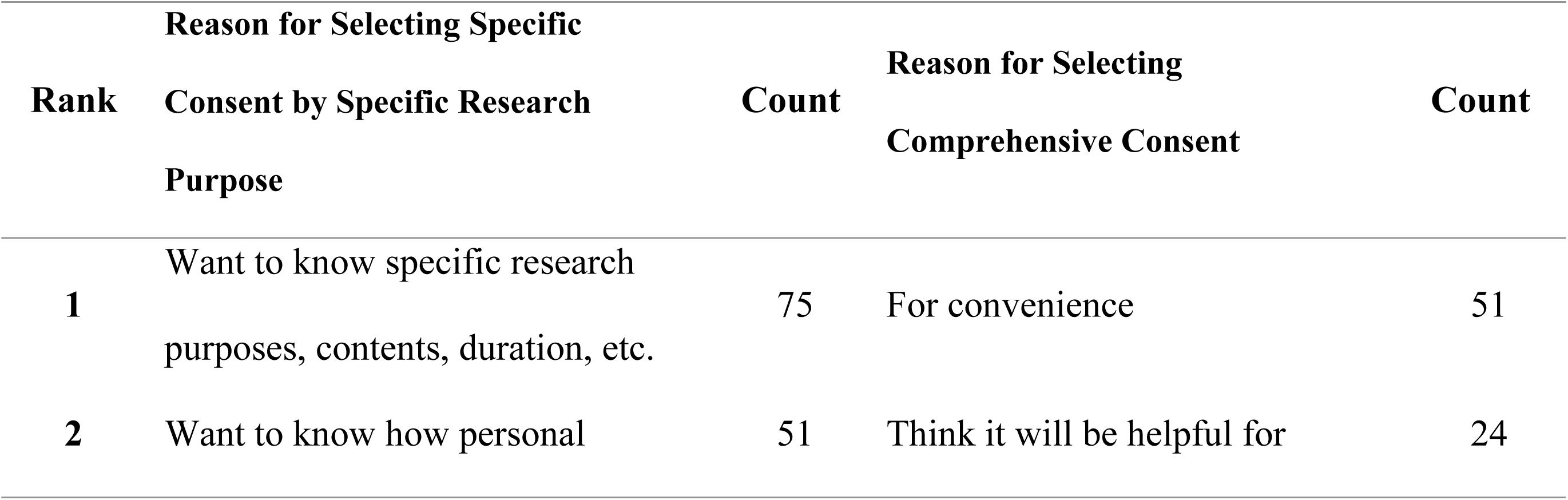
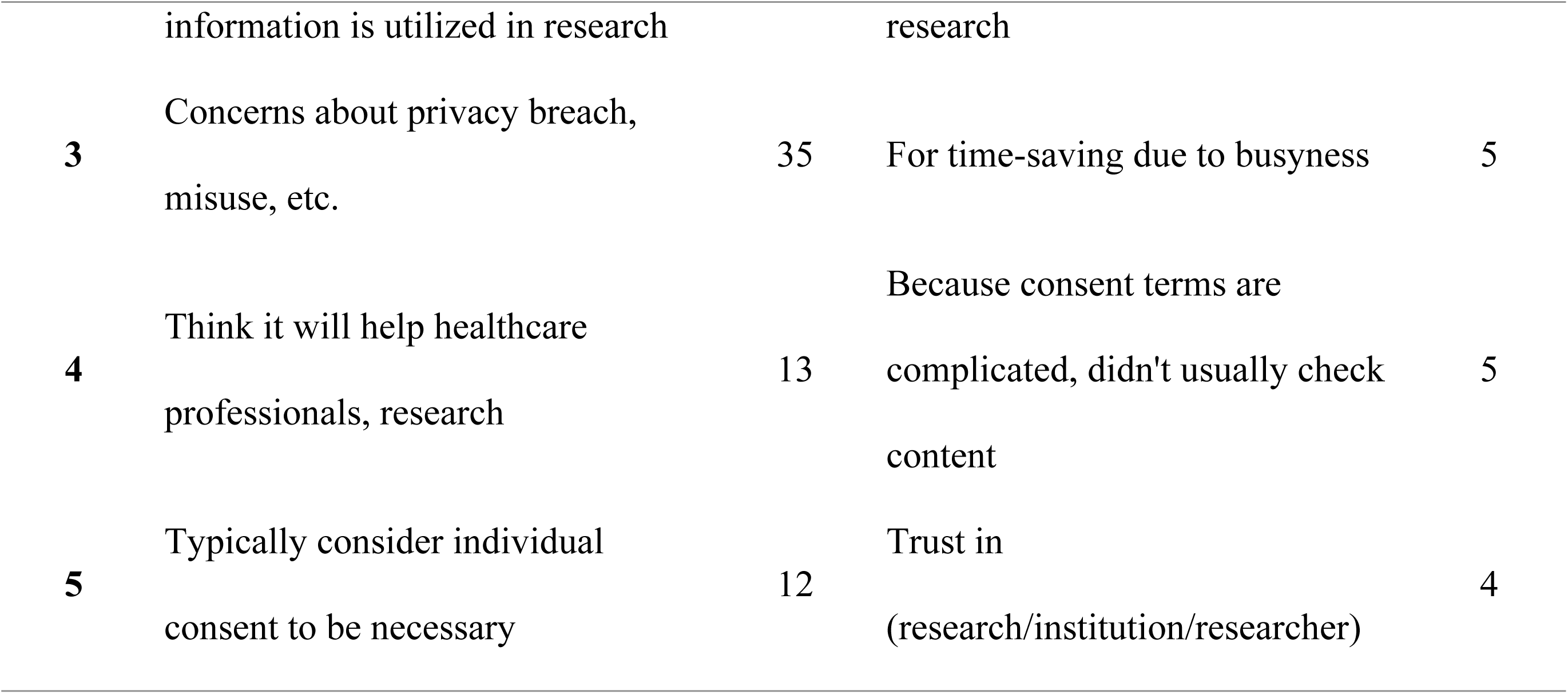
Comparison of Reasons for Selecting Specific vs. Comprehensive Consent Among Respondents.

The order in which respondents chose to answer the question “What are the most important steps or information you would like to know when consenting to the use of your/your family’s PGHD?” was as follows: 1. Results of data analysis, research results, and implications (41.5%, 166/400); 2. Data protection and security issues related to data utilization (25.8%, 103/400); 3. Contact information on the person in charge (17.5%, 70/400); 4. How to withdraw consent (14.5%, 58/400); 5. Others (0.7%, 3/400). All groups except the 20s respondent and traumatic conditions groups responded that they most wanted to know about the results of PGHD utilization. These groups selected data protection and security issues as their top priority.

### Other sub-responses and responses to open-ended questions

Among the other responses regarding “Methods used to record or manage your/your family’s main health problems (A5a),” there were responses indicating the utilization of domestic, photo-centric social networking service (SNS) applications, blogs and KakaoStory.(19) Additionally, there were responses indicating management through devices capable of measuring blood glucose levels.

Among the content collected from open-ended and other responses, when asked about the best aspects of using tethered PHR for managing or accessing health information (B2a1), responses included statements such as “It is easier to check test results compared to printed documents” and “I can directly access test results without having to print documents or inquire at the hospital.” Additionally, in the B2a1 question asking for additional explanations regarding the selected responses, respondents who chose “① It was helpful in the self-management of health and diseases” provided additional opinions such as “I can thoroughly review test results” and “It is beneficial to be able to double-check test results and appointment details.” Among respondents who selected “② I have a better understanding of the healthcare provider’s explanations or treatment plans,” opinions such as “Studying and reviewing test results before appointments greatly helps in understanding, allowing for more in-depth questions to be asked to the attending physician,” were expressed. Additionally, there were statements like, “It’s great to be able to inquire directly with the attending physician about any questions I have after reviewing test results, enabling a better consultation experience.”

Among respondents who had experience managing or seeking health information through patient apps, when asked about the frustrating aspect of using the app (B2a2), one of the other opinions expressed was “I wish test results could accumulate for over a year.” Additionally, those who selected the response “② I was rather confused because I did not know which information was correct” expressed opinions such as “I felt disappointed when occasionally there were readings that raised suspicion of measurement errors” and “It would be helpful if easily understandable terms were used.”

## Discussion

This cross-sectional survey examined whether there were significant difference in the patient or caregiver patterns of health information management and attitudes toward PGHD usage, in accordance with health-related roles, disease types, and age group.

### Principal Results

We observed differences between health-related role respondent groups based on whether they were patients themselves or guardians of patients who are minors, as well as their age and gender demographics. There were differences in the experience and frequency of recording PGHD among the health-related groups. Patients themselves and guardians of minor patients (CG-minor) showed higher rates of PGHD recording and daily usage compared to guardians of elderly or other adult patients. And we found that various types of patients and their guardians were favorable toward the use of PGHD (average agreement rate of 82.9%), with the most common reason for not agreeing with, or having concerns about, its use being personal information leakage.

Research on the user-specific characteristics of PGHD services, particularly with regard to the patients themselves and their guardians, remains sparse. A prior study reported that elderly adopters (mean age 51.81 vs 43.81 years) tended to use the PGHD functions continuously.(20) However, that previous report compared two groups without specifically delineating between age groups nor differentiating between older adults directly utilizing the services and guardians doing so on their behalf and recording the data. We found herein that the PGHD recording experience significantly differed by age and disease (*P* < 0.01), and that the chronic disease group had the highest usage rate of PGHD recording experience across the categories of disease type, health role, and age group (78.8%, 134/170). The next highest rate was among the guardians of minor patients (CG-minor) (75.0%, 69/92). These findings suggest that other than clinical needs requiring continuous information management, health-related roles may also affect the active use of PGHD.

The significance of our present study findings lies in the fact that we evaluated whether the actual users were patients themselves or their guardians and analyzed this distinction to derive meaningful results. Moreover, significant differences were observed in the utilization of patient apps provided by healthcare organizations, with guardians of minor patients exhibiting a higher willingness to do so. Regarding consent for data usage, guardians of minor patients were more inclined towards comprehensive consent including identifiable data, while guardians in other groups preferred anonymous data inclusion.

These observations are related to the survey results showing that in cases where the response was ‘strongly agree’ to the use of PGHD, more people used it for research purposes (n=100) than for clinical purposes (n=95). It seems that the subjects responded more actively to the use of PGHD for research purposes, even though it is a secondary purpose, because it has the potential to solve problems faced by cancer patients or patients with rare diseases. The respondents with the highest percentage of ‘strongly agree’ responses to the use of PGHD for research purposes were the malignant disease group (33.1%, 40/121), and the group with the highest percentage of responses favoring research purposes over clinical purposes was the congenital/genetic disease group, with 28.6% (14/49) selecting research purposes compared to 18.4% (9/49) selecting clinical purposes, showing a 10.2% difference.

Our current analysis revealed disparities across age groups. Younger adults (20s and 30s) exhibited lower rates of PGHD recording compared to the 40s and over 50 group. Additionally, the reasons given for discontinuing health management app usage differed by age. Younger adults cited annoyance as the primary reason, whereas older adults reported difficulty in using the apps. Furthermore, younger adults displayed a lower intention to use such services compared to those in their 40s and over 50. Notably, young adults (20s) also showed a tendency to be less agreeable towards the research purpose utilization of PGHD. The 20s group chose data protection and security issues as the information they most wanted to know when agreeing to allow the use of their PGHD.

Consistent with these findings, a prior study has demonstrated that younger users expressed stronger privacy concerns regarding mobile health (mHealth) services compared to older users.(21) This aligns with the established notion that demographic characteristics influence behavioral intentions among potential users.(22) However, age differences in the context of PGHD use remains an under-researched area. Our present findings and those of prior reports collectively underscore the differences in attitudes and experiences towards health information recording and management among patients and guardians, as well as across different age groups. Based on these results, the provision of personalized health information recording and management services could enhance participation from each group. Moreover, when conducting research or developing services related to PGHD utilization, consideration of the unique characteristics of each group will be crucial to promote diverse participation in healthcare.

Regarding consent for the secondary use of PGHD, the most common reason for choosing specific (individual) consent was a better understanding of the use process. The finding that ‘Want to know specific research purposes, contents, duration, etc.’ and ‘Want to know how personal information is utilized in research’ were the highest-ranking responses indicated that transparency in data use and sharing of the use procedures with data providers will be important drivers of the active participation of PGHD use. In addition, ‘For convenience’ was the most frequent reason given for choosing comprehensive consent, which suggested that regardless of the consent method applied, the most important priority is sufficient understanding by the information provider (patient, guardian) as well as simplicity of the procedures. This will be a crucial reference point going forward if developing a system or application to obtain consent from patients or guardians for the secondary use of PGHD.

### Limitations

The biggest limitation of this study was that it was conducted as an online survey and the target audience comprised individuals who were participating in the patient community, which may not represent the general patient and caregiver population due to potential bias in interest or engagement. The survey of only 400 respondents also cannot represent the views of all patients or their guardians. Online surveys are limited but the fact that participants will more than likely be able to easily access the Internet, and people who participate in the patient community are likely to have more active tendencies than patients or guardians who do not.(23) According to the 2016 National Statistics of Korea, the median outpatient age was 50-54 years old, and about one-third (31.6%) of the total were 65 years or older.(24) On the other hand, the age group with the most participants in this survey was the 40s (n=170), and those aged 50 or older accounted for only 14.5% (58/400) of the total, which is younger than the age distribution of patients estimated in 2016. In addition, there is a limitation that pediatric/elderly patients could not directly respond.

Notwithstanding these drawbacks, the respondents were likely to have high accessibility to online information and be in a position to make decisions about the use of PGHD. Hence, although bias in our selection of respondents was inevitable, the results of this study can be said to better reflect the real world.

This study was based on the results of a survey conducted only in Korea, so the results may not be generalizable. However, Korea has a high level of development and maturity in EMR, and its citizens have high accessibility to online information, so it was a meaningful cohort to use to start this discussion.(25, 26) In addition, a national project in Korea for patient-centered health data utilization is being promoted, so it is an important time to confirm the awareness of PGHD utilization and reflect it in the establishment of systems and processes.(27) This survey confirmed patient awareness of PGHD usage and utilization, but was limited by its cross-sectional design. People’s awareness can change as their health status worsens or improves, and health-related roles are also dynamic over time. Hence, the questionnaire was limited in its ability to address patient expectations and concerns. Additionally, technological advancements and policy changes can also affect patient awareness or attitudes towards PGHD, so a continuous, multidisciplinary approach and further research is needed.

### Conclusions

A cross-sectional survey was used to examine differences in patient or caregiver patterns, attitudes and anxiety towards PGHD management and usage, in accordance with their life situation. Most of the patient and caregiver respondents were favorable toward the use of PGHD, but there were concerns about the security of personal information. The respondents in their 20s and suffering from traumatic conditions were the most reluctant to use PGHD, largely due to these privacy concerns.

## Data Availability

The data underlying the results presented in the study are available from the Asan Medical Center Institutional Data Access / Ethics Committee. However, public disclosure of the data is not feasible as data sharing was not considered during the study design and IRB review. Researchers who meet the criteria for access to confidential data can contact the committee at or +82-02-3010-7166.

https://www.dropbox.com/scl/fi/pblgnljhvvcrdn4ltzb8q/Supplementary-Tables-ST1-2-3.pdf?rlkey=ekpoc0ia424t8p80lj2h3cnov&st=s074386x&dl=0

## Acknowledgements

This research was funded by the Asan Institute of Life Sciences, Asan Medical Center, Seoul, Korea (grant number 2022IP0067-2).

## Author Contributions

**Conceptualization**: Yura Lee, Sang Sook Beck

**Data curation**: Ye-Eun Park

**Formal analysis**: Ye-Eun Park, Yura Lee

**Funding acquisition**: Yura Lee

**Investigation**: Sang Sook Beck, Ye-Eun Park

**Methodology**: Yura Lee

**Project administration**: Yura Lee

**Resources**: Ye-Eun Park, Sang Sook Beck, Yura Lee

**Supervision**: Yura Lee

**Validation**: Ye-Eun Park, Yura Lee

**Visualization**: Ye-Eun Park

**Writing – original draft**: Sang Sook Beck, Ye-Eun Park

**Writing – review & editing**: Ye-Eun Park, Yura Lee

## Abbreviations

CG-adult: caregivers of adult patients not falling into the previous categories
CG-minor: caregivers of minor patients
CG-elder: caregivers of patients aged 70 or older or those unable to attend medical appointments alone
PGHD: Patient-generated health data
Pt: patients themselves
PHR: Personal Health Record

